# The impact of the Impella RP^®^ device on a failing right heart. A modelling and simulation approach to ascertain its potential

**DOI:** 10.64898/2026.06.24.26356428

**Authors:** Beatrice De Lazzari, Angela Richter, Christoph Nix, Roberto Badagliacca, Annalisa Pitino, Mercedes Gori, Gianmarco Scoccia, Massimo Capoccia, Claudio De Lazzari

## Abstract

**Background and Objective:** Indications for right ventricular assist device (RVAD) insertion include right heart failure after implantation of a left ventricular assist device or early graft failure following heart transplantation. This study aimed to investigate how the upstream and downstream circulatory network interacts with the Impella RP® device

**Methods:** A numerical model of the Impella RP® was implemented within CARDIOSIM© software platform for this study. In the numerical configuration, the RVAD aspirated blood from either the right atrium (RA-PA connection) or the right ventricle (RV-PA connection) and delivered it to the pulmonary artery. Only RA-PA connection is the currently used setting for Impella RP® in clinical practice. Based on right ventricular (RV) decompression and total flow, our study may help define the need for a direct RV-unloading Impella RP®

**Results:** The simulations showed that activating the RVAD in RA-PA mode, regardless of its rotational speed, the mean pulmonary artery pressure (PAP) percentage change was higher than the unsupported condition when the mean systemic venous pressure (SVP) and the pulmonary artery wedge pressure (PAWP) were both set to 20 mmHg. When RV-PA connection was applied, a similar trend was observed although the PAP percentage changes were about halved compared to the RA-PA connection

**Conclusions:** The Impella RP^®^ has the potential to become a valid option for RV support based on current experimental and simulation data. Although already in use, further evaluation in the clinical setting will likely confirm its potential and lead to a more routinely application for RV support.

## Introduction

The prognostic role of the right ventricle (RV) should be considered in the context of left-sided heart failure [1]. Failure of systolic function adaptation leads to increased dimensions with a negative effect on diastolic ventricular interactions [2]. RV-PA coupling has significant reserve in the context of elevated RV afterload, although the level of uncoupling that leads to RV failure remains not completely defined [3]. Better understanding of the pathophysiology of RV failure may well help with its initial medical management and timing of mechanical circulatory support. Prolonged survival by effective medical treatment becomes the grounds for the development of right heart failure secondary to chronic left ventricular dysfunction. Patients remain compensated as long as the right ventricle is functional. The ability to track the RV based on better monitoring of afterload and functional reserve may help change the course of the disease before the RV reaches the threshold that may limit both medical and LVAD treatment [4]. Right ventricular failure remains quite a challenging condition to manage in view of its complex background and still incomplete understanding of its pathophysiology [5–7]. Indications for right ventricular assist device (RVAD) insertion include right heart failure after implantation of a left ventricular assist device (LVAD) or early graft failure following heart transplantation. Right ventricular pathophysiology in the setting of pulmonary hypertension [3] represents the prevalent pathological condition for RVAD insertion, whether to support a failing left ventricle or a primarily involved right ventricle [1]. The development of RV dysfunction in end-stage lung disease is related to acute or chronic RV afterload increase due to the underlying lung disease or hypercapnic/hypoxic respiratory failure. Initial management of acute RV dysfunction includes volume optimisation, afterload reduction (nitric oxide), inotropes to restore contractility (milrinone, dobutamine) and improvement of systemic perfusion (noradrenaline, adrenaline, vasopressin). The presence of pulmonary hypertension would require pulmonary vasodilators such as prostacycline, phosphodiesterase-5 inhibitors and endothelin receptor antagonists. Persistence of RV dysfunction leading to failure would be an indication for extracorporeal life support (ECLS). Although veno-venous extracorporeal membrane oxygenation (VV ECMO) can be considered an initial strategy in the presence of RV dysfunction, it does not provide RV support [8–11] requiring reconfiguration to veno-pulmonary ECMO (VP ECMO) [12] using the ProtekDuo, which is a single-site dual-lumen cannula for percutaneous insertion through the right internal jugular vein and placement of its tip in the main pulmonary artery [13, 14]. The proximal fenestration of the cannula drains venous blood from the right atrium (RA) to the ECMO system followed by re-infusion into the main pulmonary artery through the distal fenestrations. This configuration, namely dual lumen (dl) V-P ECMO, bypasses the RV completely leading to suitable and effective RV assistance [15]. The use of VP ECMO may reduce the development or progression of right ventricular dysfunction and lead to a potentially successful outcome [16–19].

Percutaneous and surgically implanted right ventricular assist devices (RVADs) have been investigated in different clinical settings [20]. We have previously focused our attention on the ProtekDuo [21], which involves a complete bypass of the right ventricle leading to improved pulmonary flow, left atrial filling pressure and left ventricular preload with reduced leftward shift of the interventricular septum observed in right ventricular failure [17, 22].

Another option would be the use of the Impella 2.5 device, an axial flow pump, which is retrogradely advanced through the aortic valve and works by aspirating blood from the left ventricle to forward it directly into the ascending aorta (LVAD). Although no longer available on the market, the device has been previously studied by our group [23, 24].

The purpose of this study is to investigate the Impella RP^®^ potential for the support of a failing right ventricle and argue its role in a clinical setting. In the study, we consider these RV conditions:

- PH-adapted describing the successful right ventricular remodelling to handle increased pressure without failing, maintaining cardiac output and normal structure, usually with reversible changes;
- PH-maladapted resulting in ventricular dilatation, dysfunction, and severe damage when the right ventricle cannot sustain the elevated pressure.

Only RA-PA connection is the currently used setting for the Impella RP^®^ in a clinical context. Based on right ventricular (RV) decompression and total flow TF (sum of pump flow and right ventricular output), the present work may define the need for the development of a direct RV-unloading Impella RP^®^ device.

## Methods

The pressure-flow curves for the Impella RP**^®^** right ventricular assist device (RVAD) plotted in Fig. 1 are the starting point for the development of a numerical model of the right cardiovascular system and the Impella RP**^®^** pump to investigate their interactions and the performance of the device as RVAD.

**Figure 1.**
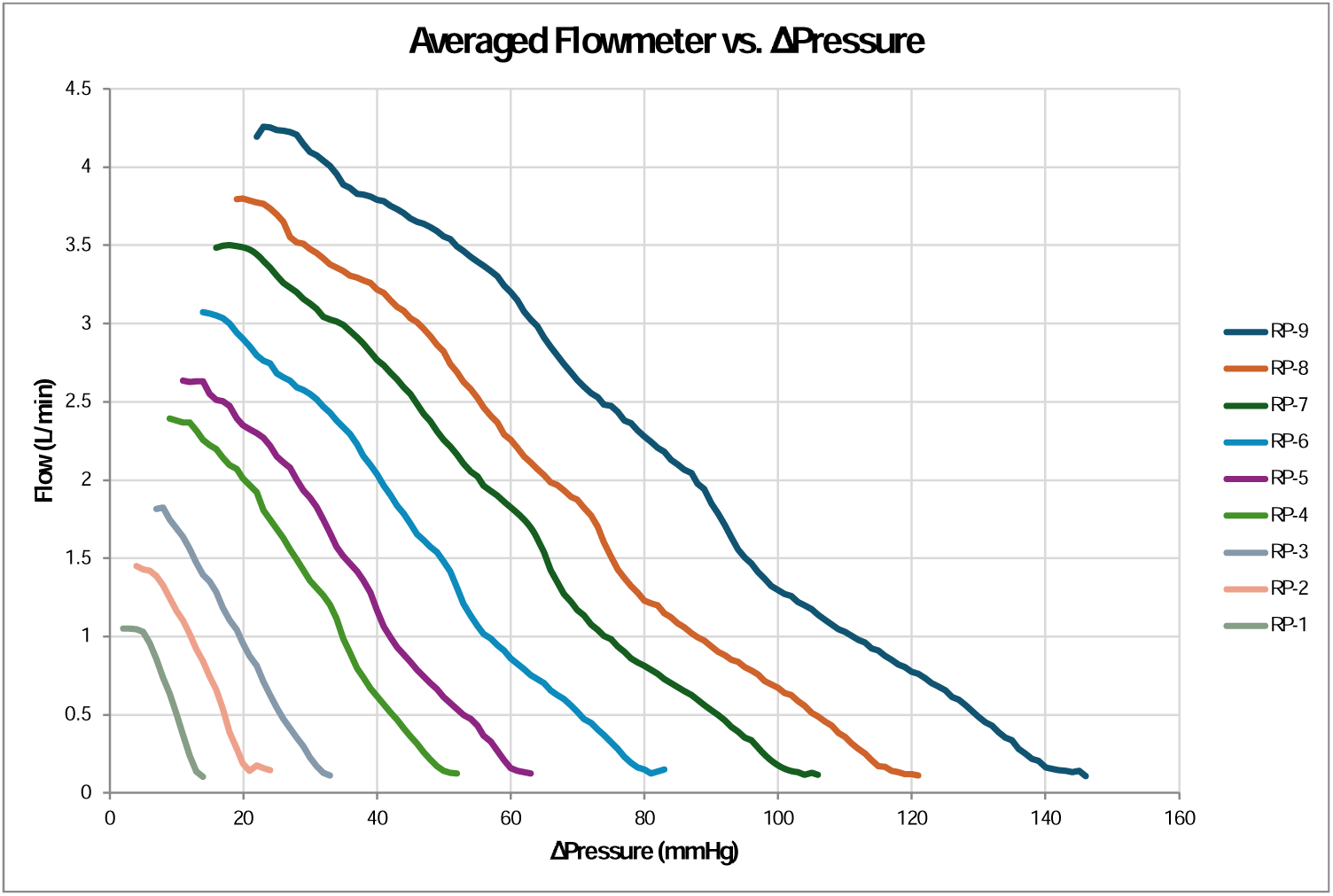
Pressure-flow curve for the Impella RP right ventricular assist device (RVAD) according to experimental data.

A new module incorporating a right circulatory network based on a time-varying elastance model was developed within CARDIOSIM^©^ software platform to replicate the behaviour of the right cardiovascular network when the Impella RP**^®^** RVAD aspirates blood from the right atrium and delivers it to the pulmonary artery (RA-PA connection) or when the RVAD aspirates blood from the right ventricle and delivers it to the pulmonary artery (RV-PA connection). In this half-circuit configuration, the right atrium and ventricle are connected to the pulmonary circulation via an RLC network.

The potential of the Impella RP**^®^** to help patients with right ventricular failure could be assessed and defined according to this arrangement by controlling the systemic filling pressure (or systemic venous pressure) (SVP) and the pulmonary artery wedge pressure (PAWP) to appropriate levels.

The following equation for the instantaneous right ventricular pressure has been used to replicate the behaviour of the right ventricle:

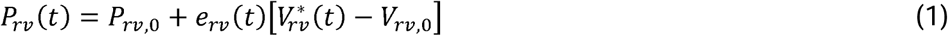

where *P_rv_(t)* is the right ventricular instantaneous pressure, *V*_rv_ (t)* is the instantaneous right ventricular free wall volume, *e_rv_(t)* is the right ventricular elastance , *V_rv,0_* is the resting right ventricular volume and *P_rv,0_* is the resting right ventricular pressure. The right ventricular elastance is described by:

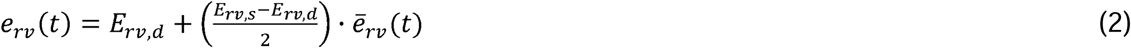

where:

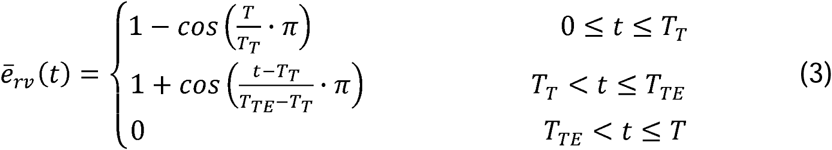

*T* is the duration of the ECG signal (Fig. 2), *T_TE_* is the end of ventricular systole and *T_T_* is the T-wave peak time. *E_rv,d_* and *E_rv,s_* are the right diastolic and systolic ventricular elastance, respectively.

**Figure 2.**
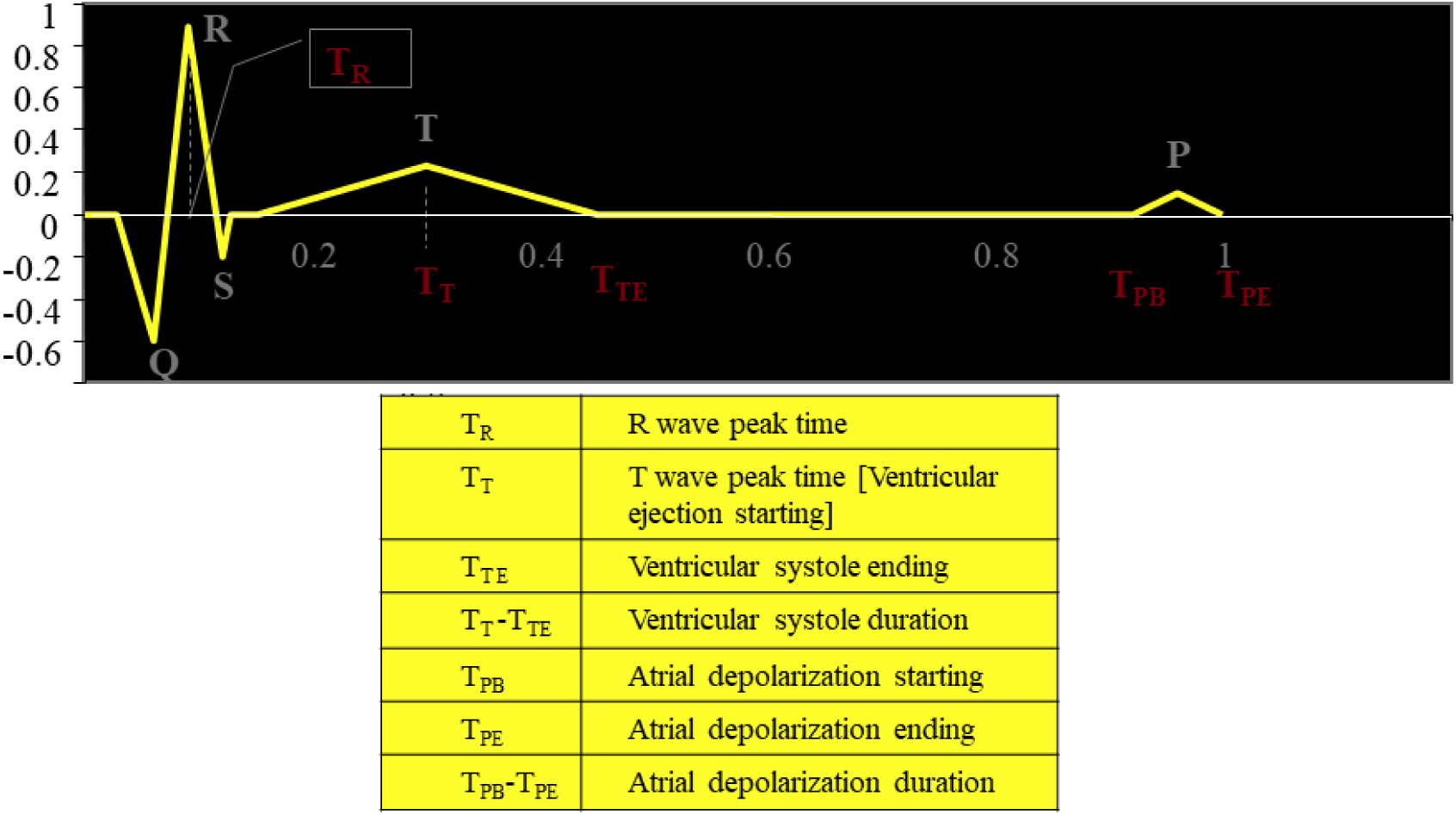
Schematic representation of the ECG signal.

The following equation for the instantaneous right atrial pressure has been used to replicate the behaviour of the right atrium:

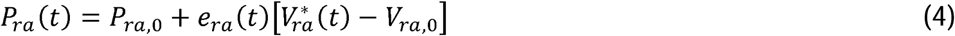

*Pra(t)* is the instantaneous right atrial pressure, [*V*ra(t)*] is the free wall volume, *e_ra_* is the right atrial elastance, *V_ra,0_*is the resting right atrial volume and *P_ra,0_* is the resting right atrial pressure.

The right atrial elastance is:

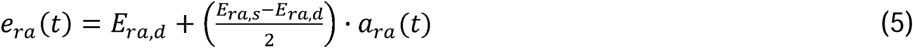

The activation function (*a_ra_* (*t*)) describing the contraction and the relaxation phases is:

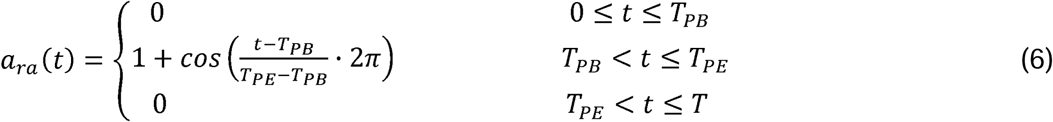

*E_ra,d_* and *E_ra,s_* are the right diastolic and systolic atrial elastance, respectively. According to the ECG, atrial systole is defined as the difference between the onset of atrial depolarization (*T_PB_*) and the end of atrial depolarization (*T_PE_*).

Figure 3 shows the electrical analogue for the new right cardio-circulatory module within CARDIOSIM^©^ platform. The picture reproduces the schematic representation of the Impella RP**^®^** used with RA-PA and RV-PA connection.

**Figure 3.**
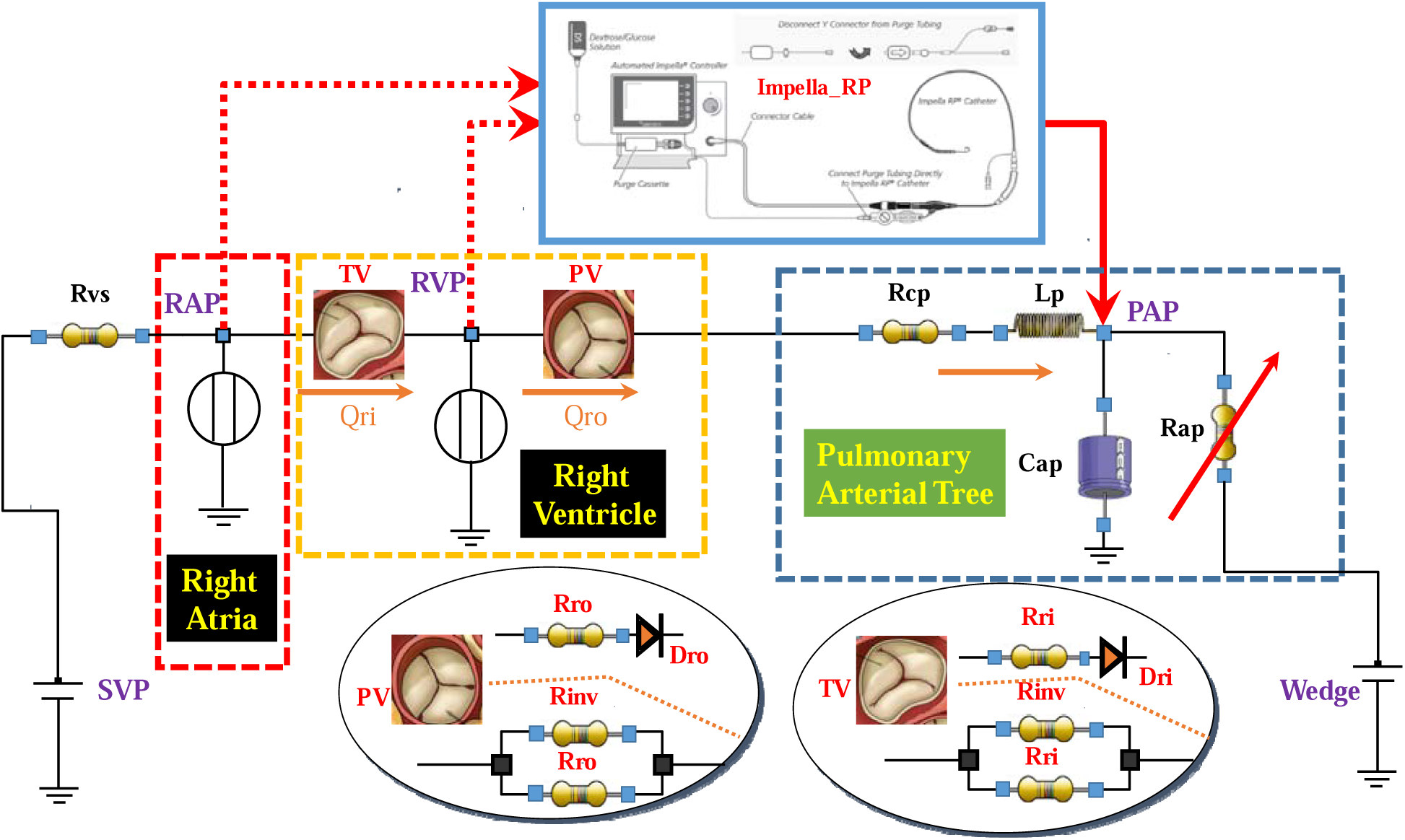
Open network that mimics the right cardiovascular system. The whole pulmonary circulatory system can be simulated using pulmonary characteristic resistance (Rcp), pulmonary arterial inductance (Lp), pulmonary arterial compliance (Cap), and varying pulmonary arterial resistance (Rap). Mean pulmonary arterial pressure (PAP), mean systemic venous pressure (SVP) and pulmonary artery wedge pressure (PAWP) can be set to constant settings.

We used the following quadratic equation to replicate the Impella RP flow:

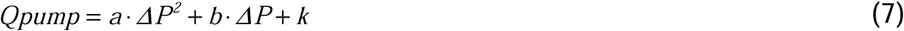

where: *ΔP* is the pressure drop (Ventricular/Pump or Atrium/Pump), *Q_pump_*is the flow produced by Impella RP^®^ and finally, *a*, *b*, and *k* are reported in the following Table 1:

**Table 1.**
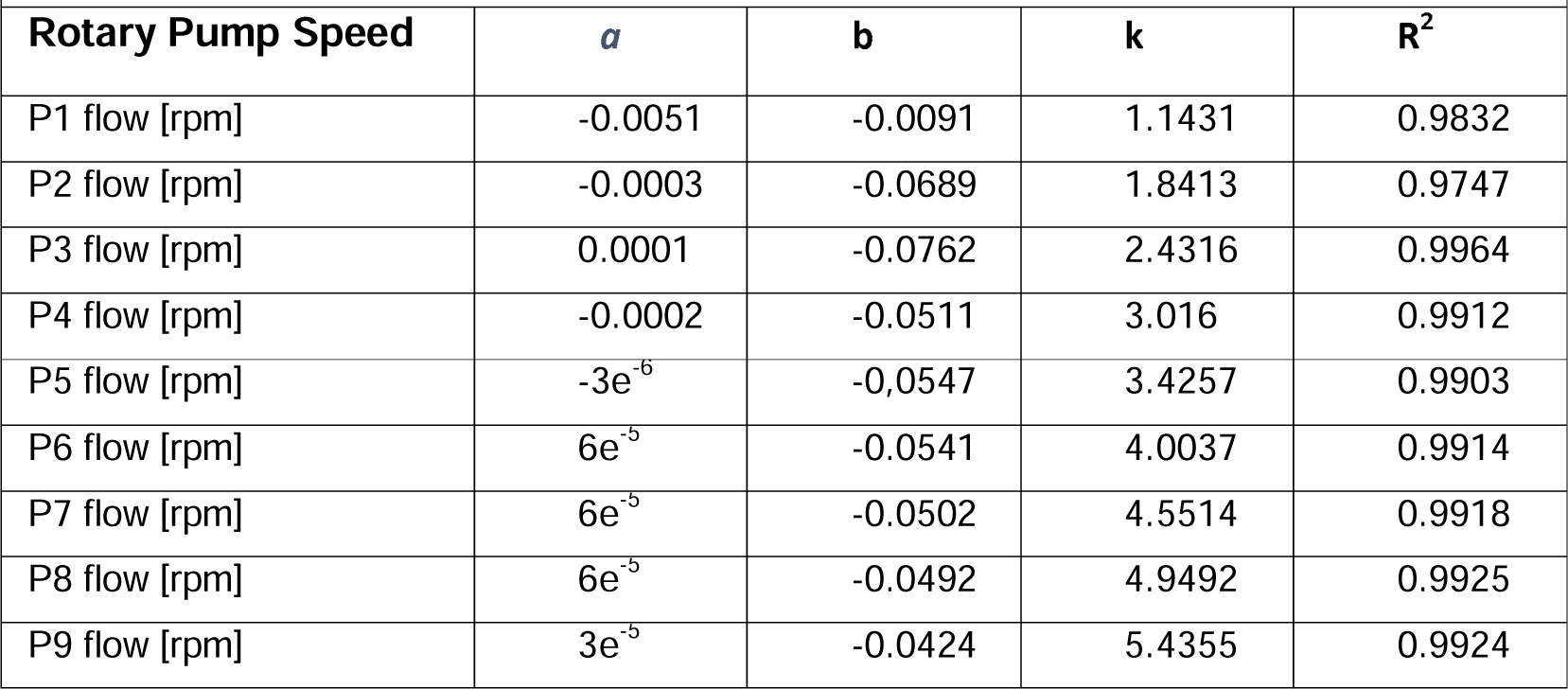
Parameters used in Eq.7.

Figures 4, 5 and 6 show all the Pressure-Flow waveforms obtained using a quadratic equation (Eq. 7):

**Figure 4.**
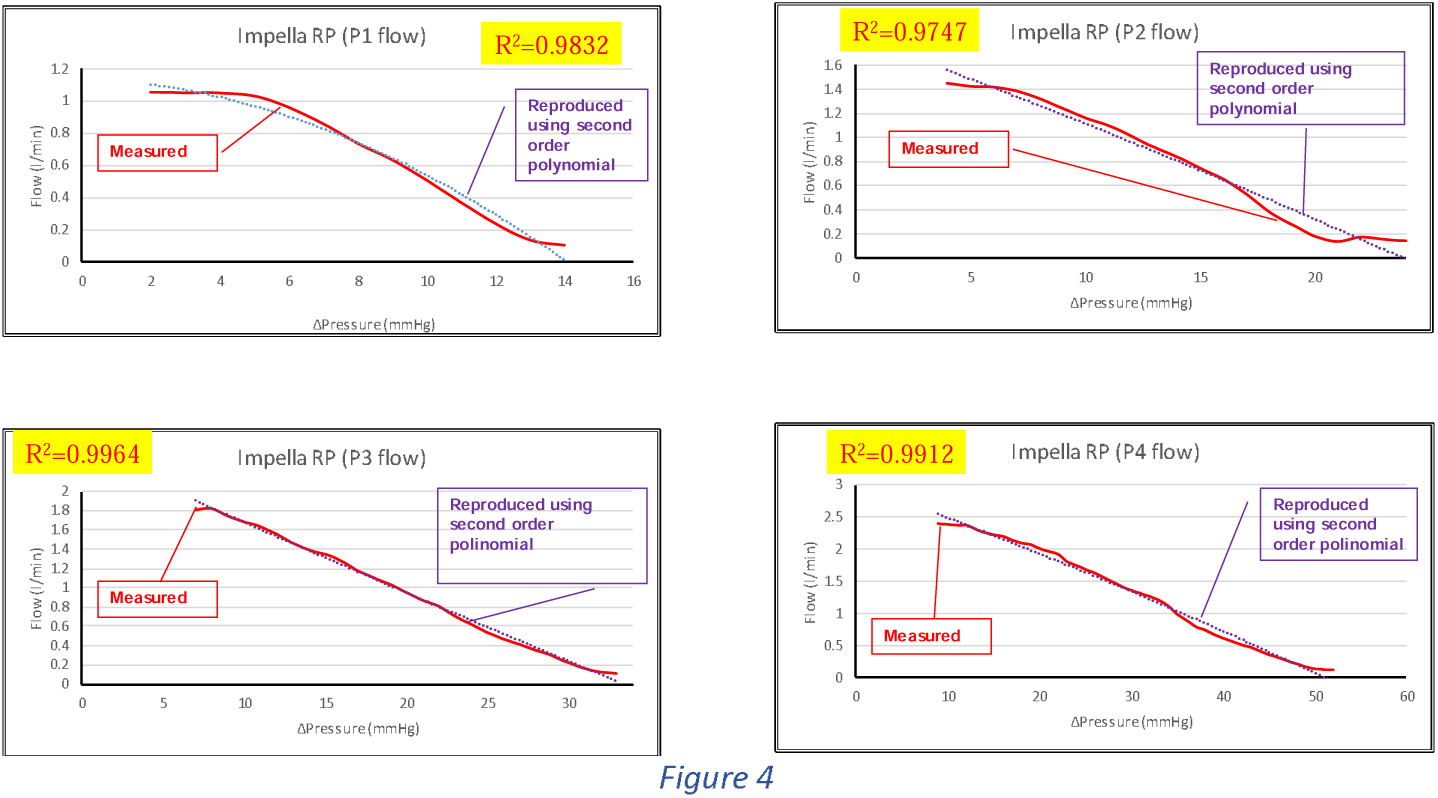

The configuration presented in Fig. 3 allows changes in the mean systemic filling pressure (SVP), the pulmonary artery wedge pressure (PAWP) and, if necessary, the afterload in terms of pulmonary artery pressure (PAP). The afterload can be set to a specific value by activating an algorithm that maintains the mean PAP at the specified value by adjusting the pulmonary arterial resistance.

Setting the SVP and contractility with activation of the Impella RP**^®^**, it is possible to assess the effects on the right ventricular output flow rate, ventricular end systolic volume (ESV), end diastolic volume (EDV), Total Flow (TF) (sum of pump flow and right ventricular output) and PAP on the pulmonary arterial branch.

Setting the SVP and afterload and changing ventricular contractility allows the evaluation of the impact of changes in the rotational speed of the Impella RP**^®^** on the right ventricular output flow rate, ESV, EDV and TF. These simulations allow the assessment of the Impella RP**^®^** in different circulatory conditions either when the RVAD is applied with RV-PA or RA-PA connection. In addition, the PAWP can be set to a constant value.

We considered the following steps to investigate the effects of the Impella RP**^®^** on several haemodynamic and energy parameters under different cardiovascular conditions:

- To reproduce a PH-adapted condition the afterload (PAP) and mean systemic venous pressure (SVP) was set at 60 and 20 mmHg, respectively; the PAWP was set to 20 mmHg. An algorithm that could automatically change Rap and Cap was implemented in CARDIOSIM^©^ to maintain a constant PAP at 60 mmHg. In this first step only Rap’s automated version had been switched on.
- Starting from the PH-adapted condition, the RVAD was applied with both RV-PA and RA-PA connection modes at different rotational speed to evaluate the effects induced on right ventricular end diastolic/systolic volume (EDV/ESV), TF, right ventricular external work (EW), right ventricular pressure-volume area (PVA), Ees/Ea ratio (right ventricular elastance/pulmonary arterial elastance).
- In the subsequent steps the systemic venous pressure (SVP) was set to 10 and 15 mmHg; the PAWP was set to 10 and 15 mmHg. The algorithm maintained the PAP fixed at 60 mmHg. Systemic venous resistance was set at 30 [mmHg‧cm^-3‧^sec]. Under these conditions, the Impella RP**^®^**was activated to aspirate blood at different rotational speeds from the right ventricle and atria, respectively.

Right ventricular remodelling in pulmonary hypertension consists of adaptation followed by exhaustion. In its early adaptive phase, the right ventricle responds appropriately in terms of hypertrophy and enhanced contractility to maintain forward flow and protect the left ventricle. Subsequently, persistent pressure overload overcomes the compensatory response leading to maladaptive changes in terms of right ventricular dilatation, reduced systolic function and increased right-sided pressures. Therefore, we have considered PH-adapted and PH maladapted conditions during our simulations to take into account the whole spectrum of right ventricular remodelling. The acronym PH stands for pulmonary hypertension.

## Results

To simulate a PH-adapted scenario, PAP and SVP were set to 60 and 20 mmHg, respectively, with PAWP set to 20 mmHg. An algorithm that automatically changed the Rap value was activated to keep a constant PAP at 60 mmHg. The pulmonary arterial compliance (Cap) remained at 4.8 [cm^3^ mmHg^-1^]. The right ventricular elastance also remained at 0.5 [cm^-3^LJmmHg]. The heart rate (HR) remained constant at 80 bpm. Figure 7 displays a CARDIOSIM^©^ screenshot for the PH-adapted conditions in which the right ventricular output flow is 3.65 l/min and Ees/Ea is 1.32.

**Figure 5.**
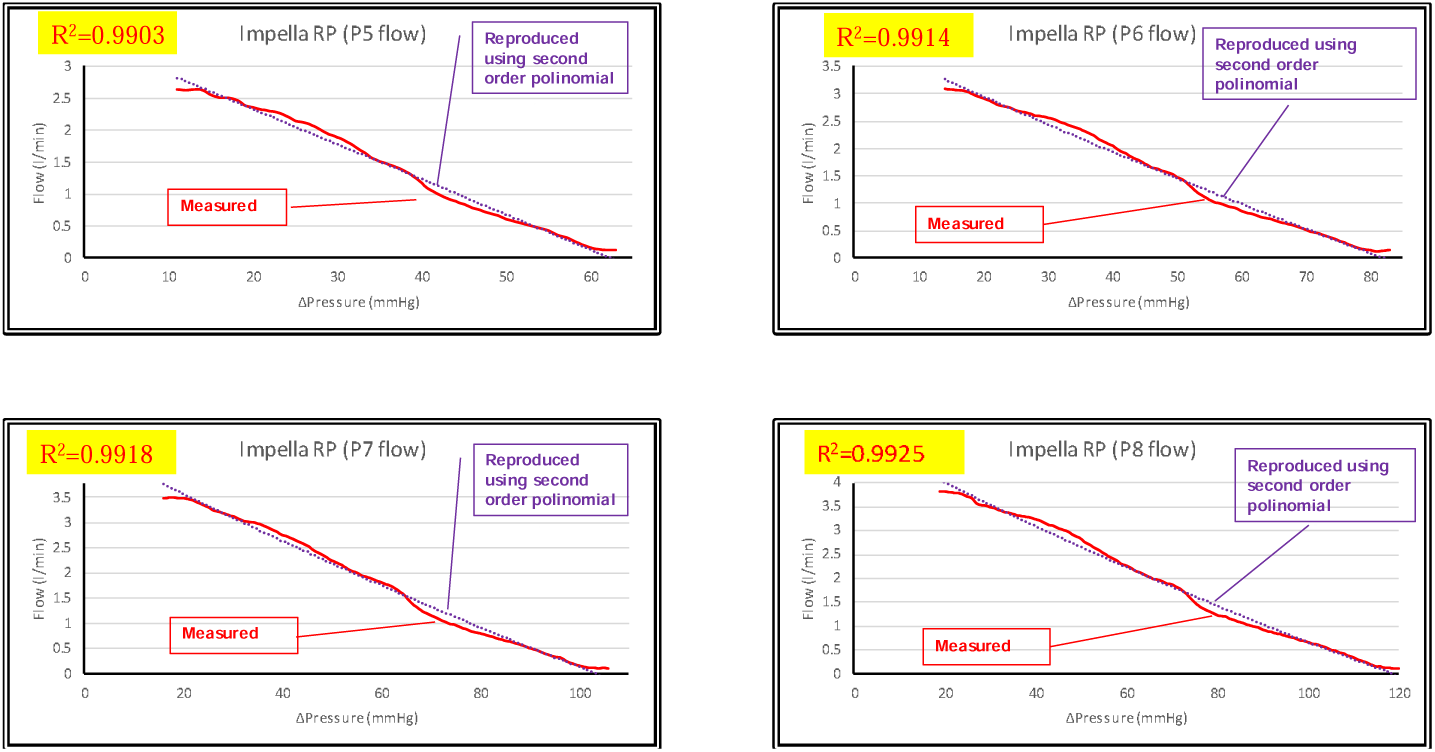

**Figure 6.**
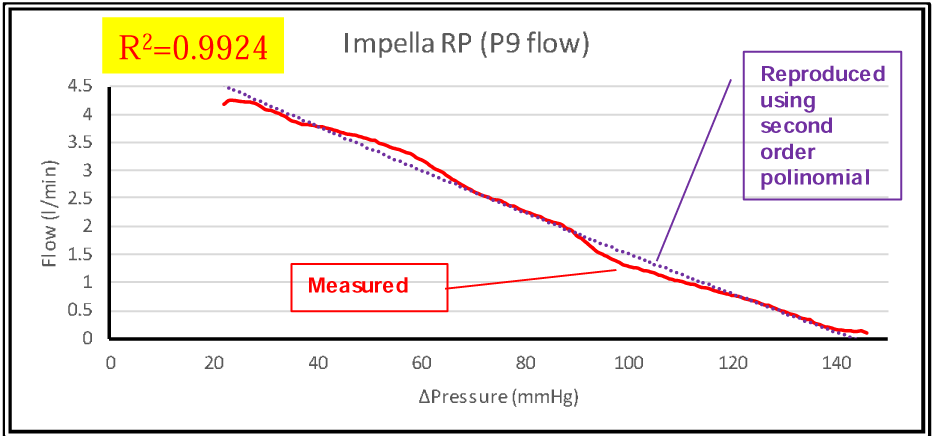

**Figure 7.**
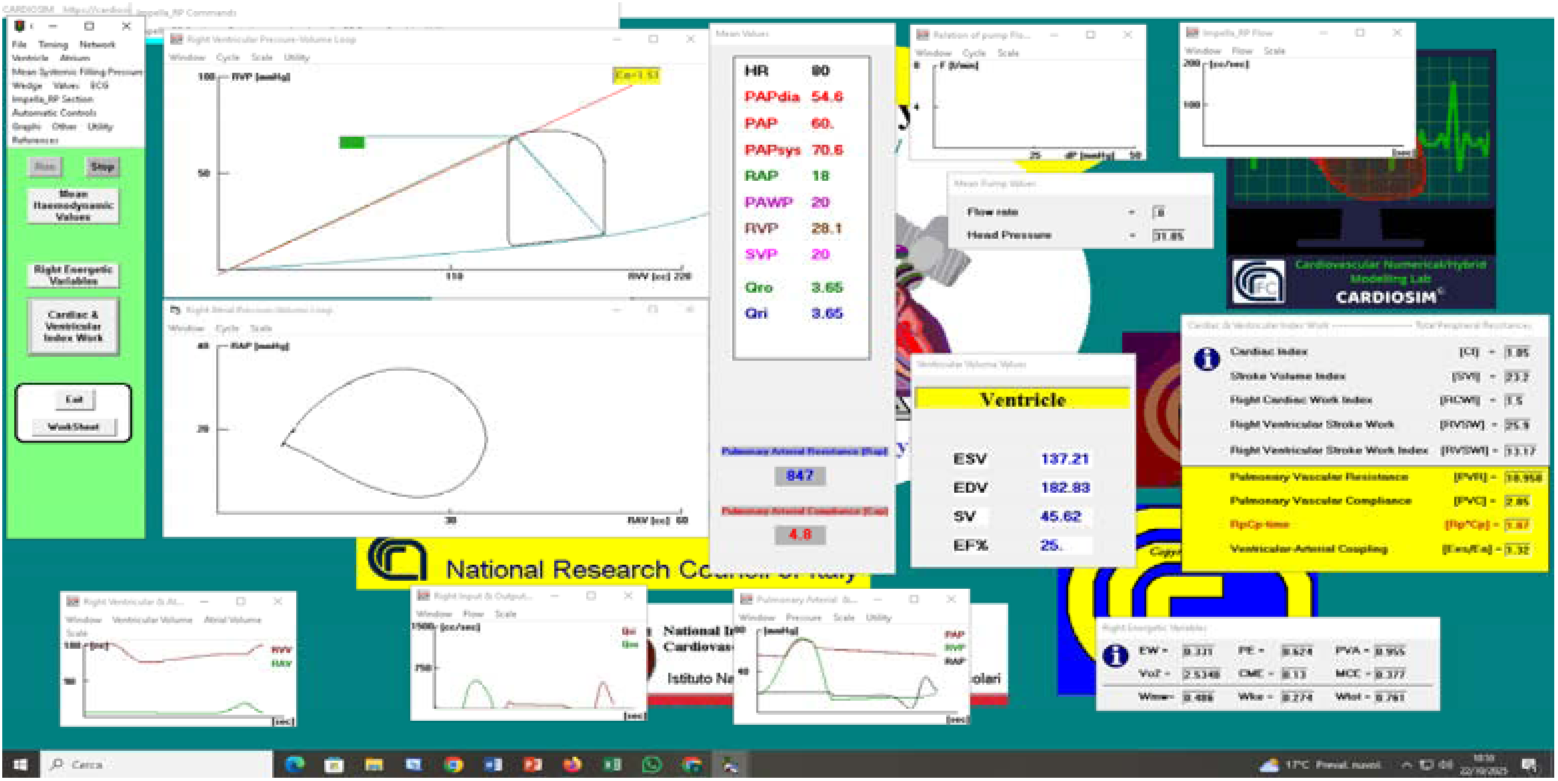
Screenshot of PH-adapted condition obtained setting the wedge to 20 mmHg. PAP was kept constant at 60 mmHg by automatically varying the Rap. Systemic venous pressure (SVP) was set to 20 mmHg. RVP is the right ventricular pressure. Qri and Qro 3 -1 are the input and output flow rates, respectively. The pulmonary arterial compliance (Cap) was set to 4.8 [cm ‧mmHg ]. The right -3 ventricular elastance was set to 0.5 [cm ‧mmHg]. HR was set to 80 bpm.

Figure 8 shows a PH-maladapted condition obtained from setting the right ventricular elastance to 0.5 [mmHg‧cm^-3^], the PAWP and SVP both to 20 mmHg, constant PAP at 60 mmHg and finally Impella RP**^®^** applied both in RV-PA and RA-PA mode with RP-5 configuration (Fig. 1).

**Figure 8.**
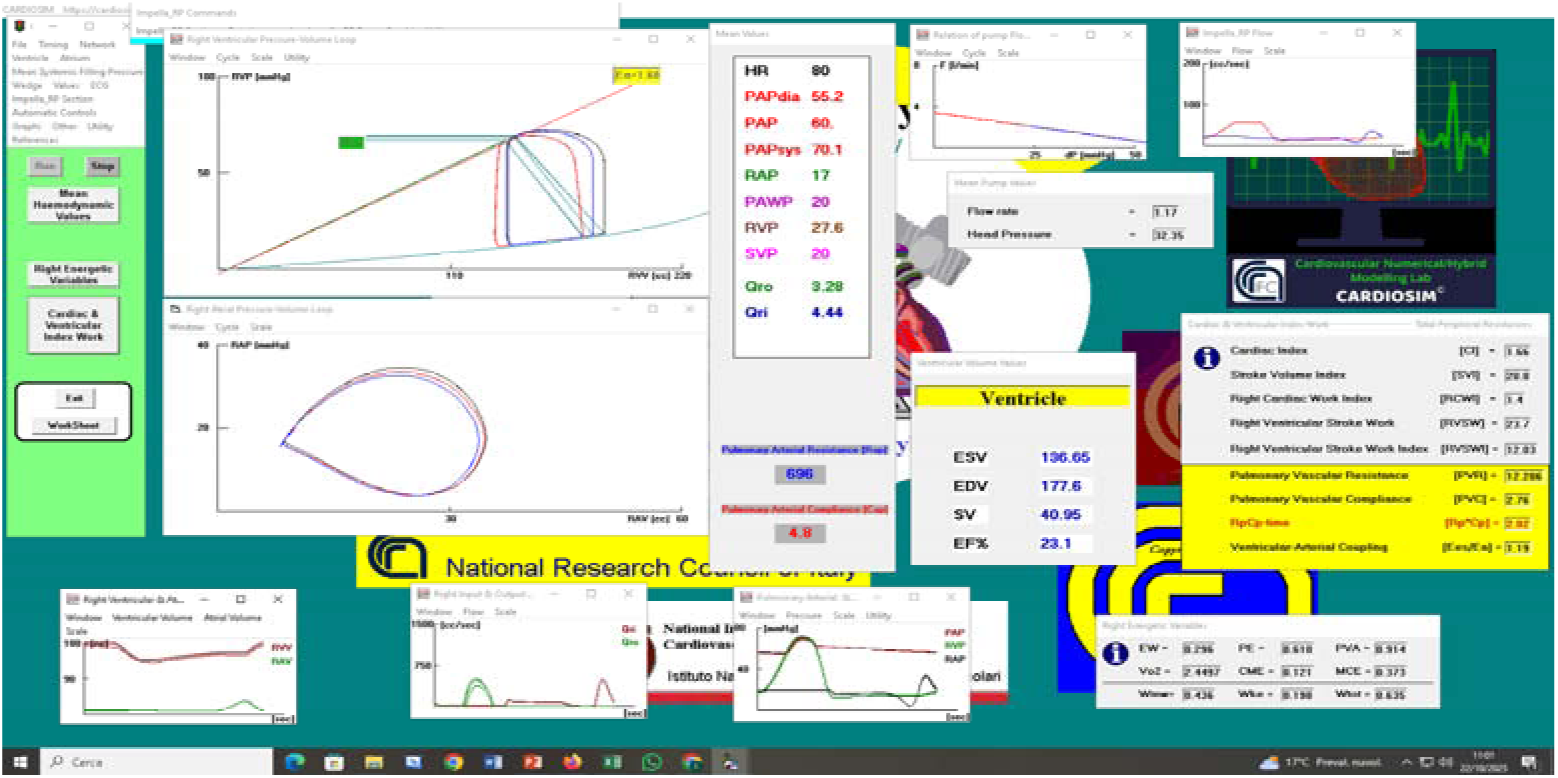
Screenshot of PH-maladapted condition setting the wedge and SVP both to 20 mmHg. PAP remained constant at 60 mmHg by automatically varying only the Rap. The right ventricular elastance was set to 0.5 [mmHg‧ cm- ]. The pulmonary arterial compliance 3 -1 ® (Cap) was set to 4.8 [cm ‧ mmHg ]. Impella RP with RP-5 rotational speed was connected in RV-PA (red loop) and RA-PA mode (blue loop). Reported mean values show the effects induced by the RVAD connected in RA-PA mode. RVP (RAP) is the right ventricular (atrial) pressure and EW (PVA) is the right ventricular external work (pressure-volume area). The RVAD flow rate was 1.59 [l/min] when Impella ® RP was applied in RV-PA mode.

The RVAD reduced the right ventricular end systolic volume (ESV) from 137.21 (Fig. 7) to 130.98 [ml] when it was applied in RV-PA mode. If the pump was connected in RA-PA mode, the ESV would be 136.65 [ml] (Fig. 8). In comparison to the unsupported condition, the EW (PVA) decreased by 10.57% (4.29%) when Impella RP**^®^**was activated in RA-PA mode and by 17.22% (10.26%) when the pump was used in RV-PA mode. The TF increased by 8.22% when the assistance was activated in RV-PA mode and by 21.64% when it was applied in RA-PA mode. When the RVAD was activated in RA-PA (RV-PA) mode, the Ees/Ea ratio decreased by 9.85% (5.30%) compared to unsupported settings.

The right ventricular end diastolic volume (EDV) decreased from 182.83 (Fig. 7) to 160.9 [ml] when the RVAD was applied in RV-PA mode with RP-7 rotational speed (Fig. 9). Alternatively, the EDV was 171.36 [ml] when the pump was attached in RA-PA mode. Impella RP**^®^** reduced the right ventricular end systolic volume (ESV) from 137.21 (Fig. 7) to 126.38 [ml] when it was applied in RV-PA mode; the RA-PA connection would give an ESV of 135.68 [ml] (Fig. 9).

**Figure 9.**
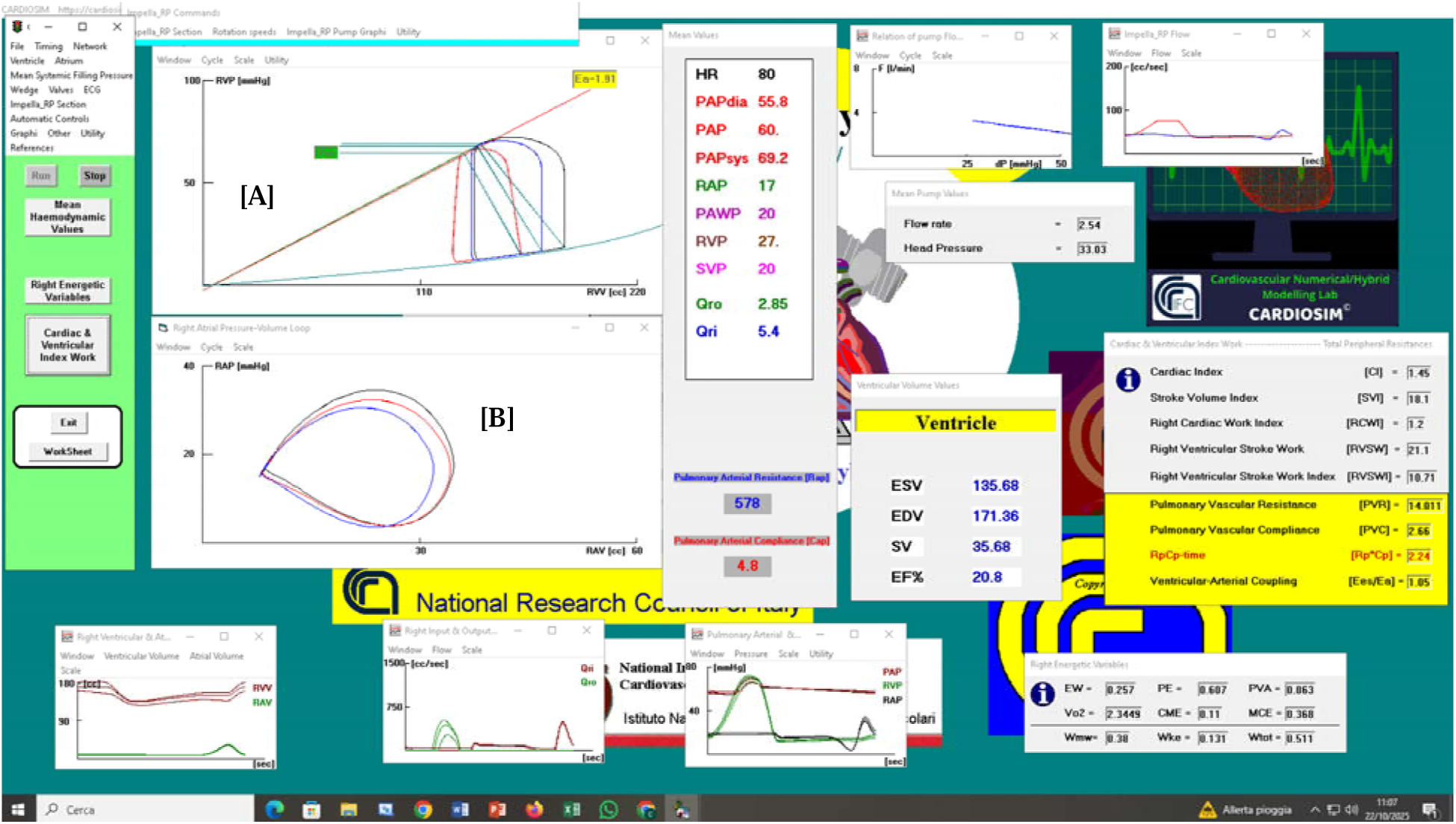
Screenshot of PH-maladapted condition setting PAWP and SVP both to 20 mmHg. PAP was kept constant at 60 mmHg by automatically varying only the Rap. The right ventricular elastance was set to 0.5 [mmHg׈cm^3^]. The pulmonary arterial compliance 3 -1 ® (Cap) was set to 4.8 [cm^−3^׈mmHg^−1^]. Impella RP with RP-7 rotational speed was connected in RV-PA (red loop) and RA-PA mode (blue loop). Reported mean values show the effects induced by the RVAD connected in RA-PA mode. The RVAD flow rate was 2.89 [l/min] ® when Impella RP was applied in RV-PA mode.

The Impella RP**^®^** flow rate was 2.54 (2.89) [l/min] when the RA-PA (RV-PA) connection was selected, whereas the TF was 5.4 (Fig. 9) and 4.28 [l/min], respectively (Fig. 9). When the RVAD was activated in RA-PA (RV-PA) mode, the Ees/Ea ratio decreased by 20.4% (18.9%) compared to unsupported settings.

In comparison to the unassisted condition, the EW (PVA) decreased by 22.36% (9.63%) when RVAD was activated in RA-PA mode and by 38.37% (21.26%) when the pump was used with RV-PA connection. When Impella RP^®^ was applied in RV-PA mode, the overall blood flow increased by 17.26%, and when it was activated in RA-PA mode, it increased by 47.95%. The right ventricular pressure-volume loop can be seen in panel [A] (Fig. 9) for both unassisted (black line) and assisted mode with the RVAD applied in RV-PA (red line) and in RA-PA (blue line) connection. Similarly, Impella RP**^®^** applied in RV-PA (red line) and in RA-PA (blue line) mode can be seen in both unassisted (black line) and assisted mode in the right atrial pressure-volume plane (loops in panel B). Impella RP**^®^** activation reduces the internal area of the atrial loop, particularly in the right atrium-pulmonary artery connection (blue line).

When the RVAD was used in RV-PA connection and activated with RP-9 rotational speed, the right EDV (ESV) dropped from 182.83 (137.2) to 150.8 (122.0) [ml] (Fig. 10).

**Figure 10.**
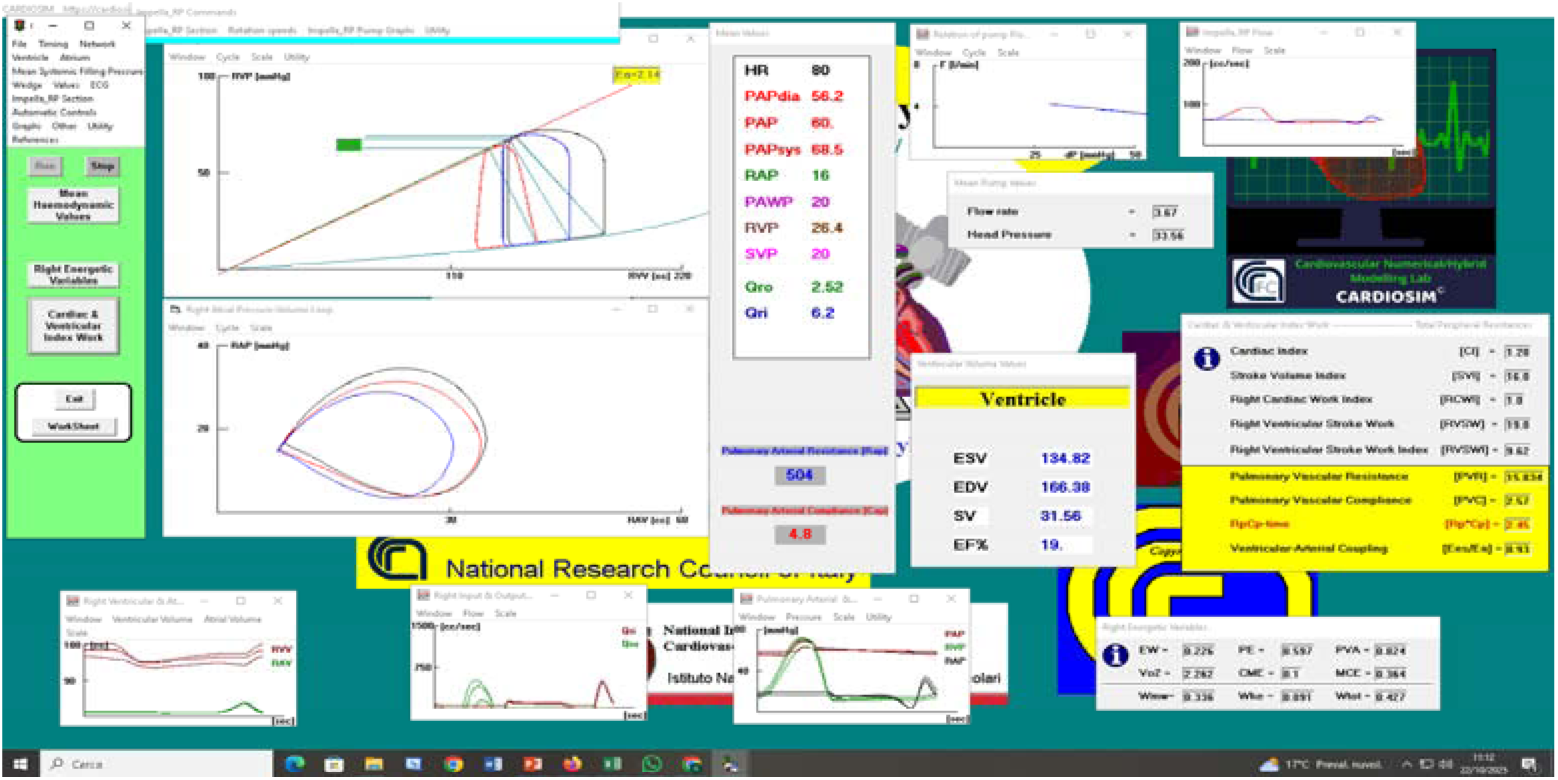
Screenshot of a PH-maladapted condition setting with Impella RP at RP-9 rotational speed. The RVAD blood flow rate was 3.95 [l/min] when applied in RV-PA mode. RV-PA (red loop) and RA-PA connection (blue loop).

The right EDV (ESV) decreased from 182.83 (137.2) to 150.82 (122.0) [ml] when the Impella RP**^®^** was applied in RV-PA mode (Fig. 10). Alternatively, the EDV (ESV) decreased to 166.38 (134.82) [ml] when the RVAD was used in RA-PA connection. When the RA-PA (RV-PA) assistance was activated, the Impella RP**^®^** flow rate was 3.67 (3.95) [l/min], while the TF was 6.2 (Fig. 10) and 4.57 [l/min], respectively.

In the following simulations SVP and PAWP were set both to 20 mmHg; PAP was not constant; systemic venous resistance was set to 60 [mmHg‧cm^-3^‧ sec]; the right ventricular elastance was set to 0.5 [mmHg‧cm^-3^] and the pulmonary arterial compliance (Cap) was set to 4.8 [cm^3^‧mmHg^-1^].

Figure 11 shows that the percentage change in mean pulmonary artery pressure (PAP) from baseline is more pronounced when the Impella RP**^®^** aspirates blood from the atrium compared to when it aspirates from the ventricle. Additionally, both right ventricular assist device (RVAD) configurations result in a decrease in the ratio Eas/Ea when mean PAP is not maintained at a constant level. Under the same conditions (Fig.11), a reduction in EDV and ESV can be observed when Impella RP**^®^** draws blood from the right ventricle (top left right panel respectively), while an increase in ESV is observed when the assist draws blood from the right atrium (top right panel), because PAP increased and the native ventricle not being able to eject.

**Figure 11.**
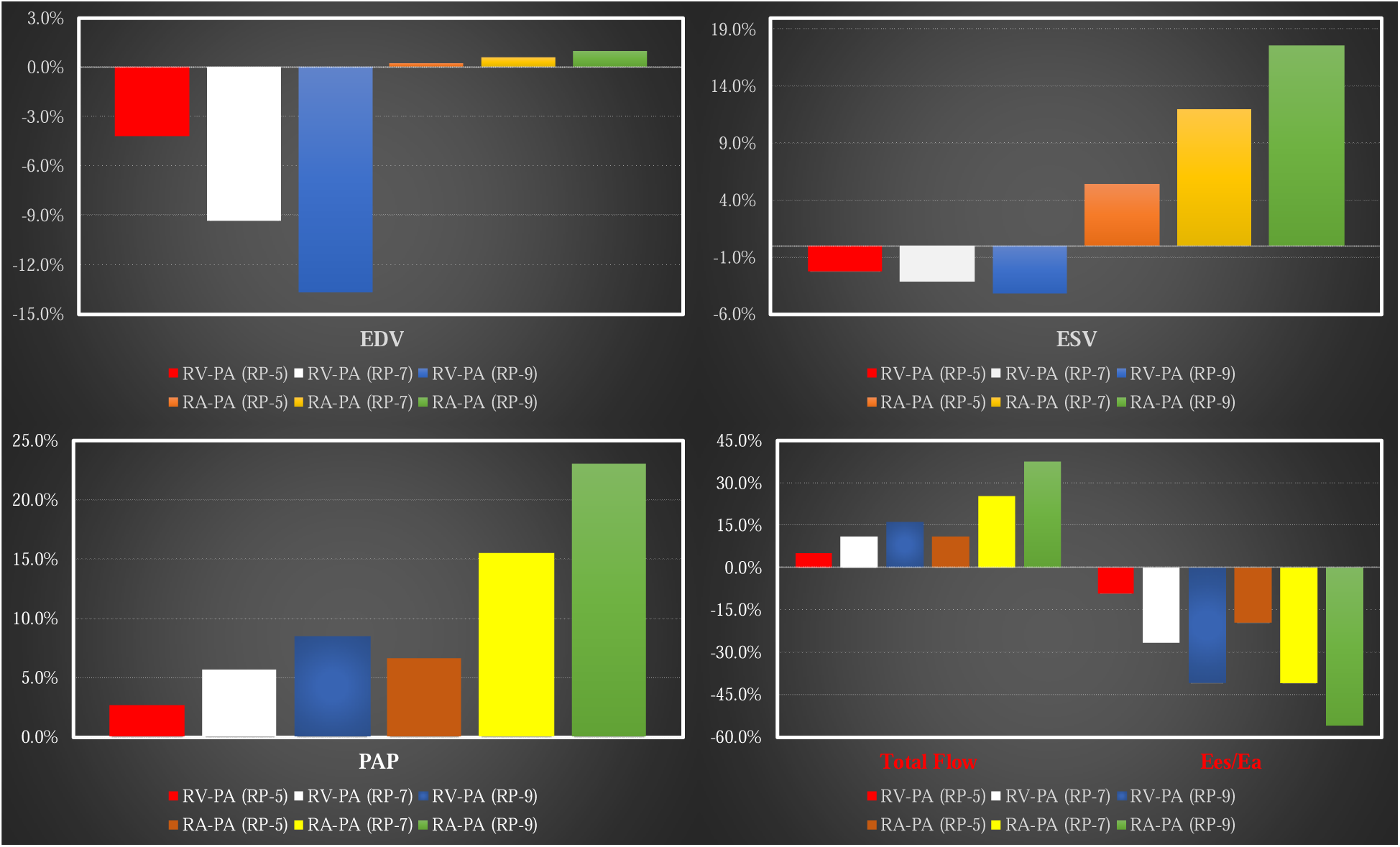
Percentage variations in right ventricular end diastolic volume (top left panel) and right end systolic volume (top right panel) in relation the following starting conditions: both systemic venous pressure (SVP) and PAWP were kept constant at 20 mmHg. Systemic venous resistance is fixed at 30 [mmHg‧cm^−3^ sec]. Bottom left (right) panel shows percentage variations in mean PAP (Total Flow and Ees/Ea).

A new set of simulations was conducted with the SVP and PAWP set to 15 mmHg, the systemic venous resistance set to 30 [mmHg cm^-3^ sec], and the mean PAP maintained at a constant value of 60 mmHg. In this case, the Impella RP**^®^** was used at different rotational speeds in both RV-PA and RA-PA configurations. Figure 12 shows the percentage variations in right ventricular end diastolic volume (right panel) and right end systolic volume (left panel) obtained when Impella RP**^®^** was activated in RV-PA and in RA-PA mode with RP-5, RP-7 and RP-9 rotational speed.

**Figure 12.**
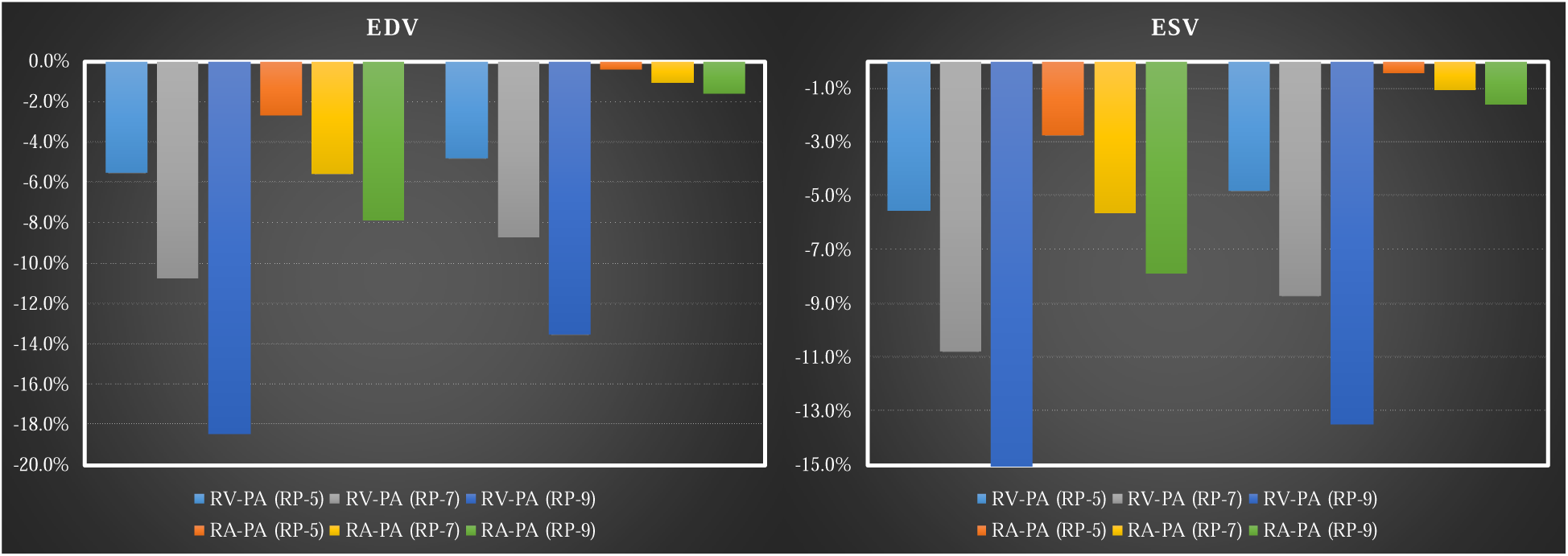
Percentage variations in right ventricular end diastolic volume (right panel) and right end systolic volume (left panel) in relation the following starting conditions: systemic venous pressure (SVP) is kept constant at 15 mmHg; the wedge set to 15 mmHg. The PAP was kept constant at 60 mmHg. Systemic venous resistance is kept constant at 30 [mmHg‧cm^−3^ sec].

Impella RP**^®^** working in both settings produces a percentage reduction in both ESV and EDV. When RVAD is connected in RV-PA mode, the percentage reduction is more evident; in this scenario, EDV reduces from approximately 6% (RP-5) to approximately 11% (RP-9), and ESV decreases from about 5% (RP-5) to approximately 14% (RP-9). There is little impact on ESV when Impella RP**^®^** is turned on in RA-PA mode at different rotational speeds (Fig. 12).

Figure 13 presents the percentage variations in TF and Ees/Ea (right panel) as well as EW and PVA (left panel) related to the starting conditions, for various activations of Impella RP**^®^** configured in RV-PA and in RA-PA modes at different rotational speeds. Higher reduction in EW and PVA is observed when the RVAD aspirates blood from the right ventricle. Under these cardiovascular settings, the ratio Ees/Ea decreases to around 29% when the RVAD (RP-9) aspirates blood from the atria and to approximately 30% when it drains blood from the ventricle.

**Figure 13.**
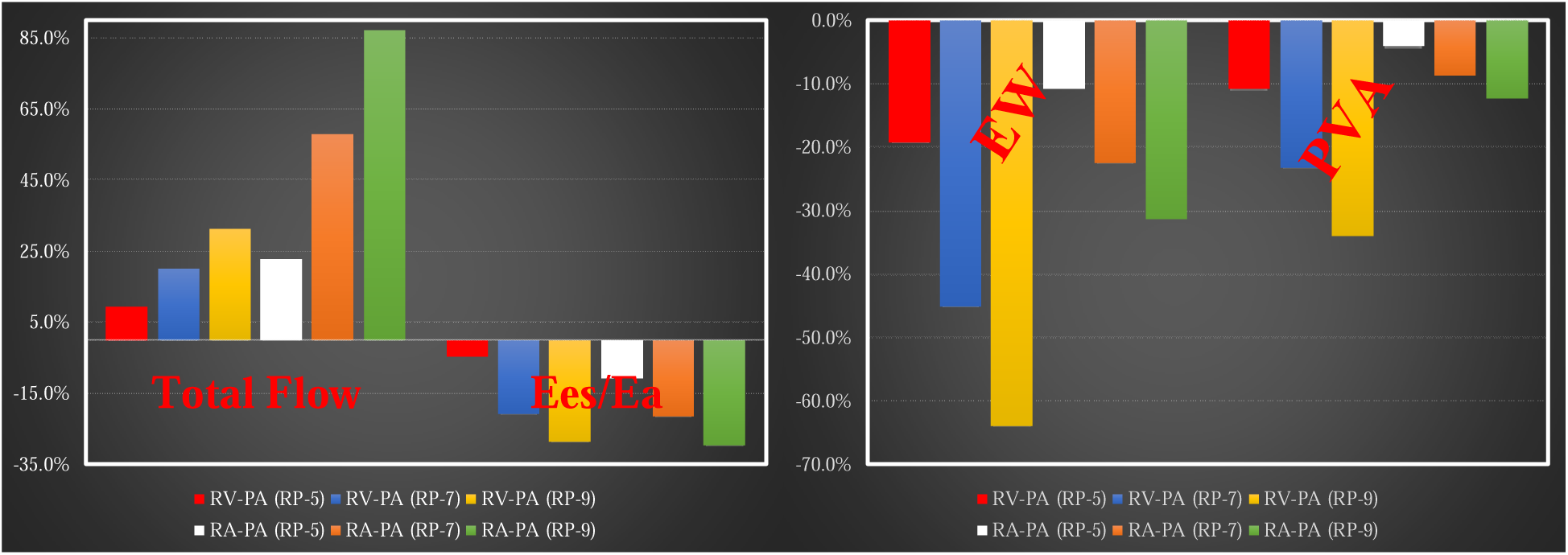
Percentage variations in TF & Ees/Ea (right panel) and EW & PVA (left panel) in relation to the following starting conditions: systemic venous pressure (SVP) is kept constant at 15 mmHg; PAWP is set to 15 mmHg. The PAP was kept constant at 60 mmHg. -3 Systemic venous resistance is fixed at 30 [mmHg‧cm^−3^ sec].

In the following simulations SVP and PAWP were both set to 10 mmHg; PAP was kept constant at 60 mmHg; systemic venous resistance was set to 30 [mmHg‧cm^-3^‧sec] (Fig. 13). In this case, the Impella RP**^®^** was used at different rotational speeds in both RV-PA and RA-PA connections.

Figure 14 shows the percentage variations in right ventricular end diastolic volume (right upper panel) and right end systolic volume (left upper panel) obtained when Impella RP**^®^** was activated in RV-PA and in RA-PA mode with RP-5, RP-7 and RP-9 rotational speed. EDV (ESV) is reduced by approximately (more than) 50% when RVAD aspirates blood from the right ventricle with RP-9 rotational speed. Left (right) lower panel shows the percentage changes of right ventricular external work and pressure volume area (TF and Ees/Ea). Both EW and PVA decrease more when the RVAD aspirates blood from the right ventricle (left lower panel). The ratio Ees/Ea drops to about 48% when the RVAD aspirates blood from the atria, but it rises to about 90% when it drains blood from the ventricle.

**Figure 14.**
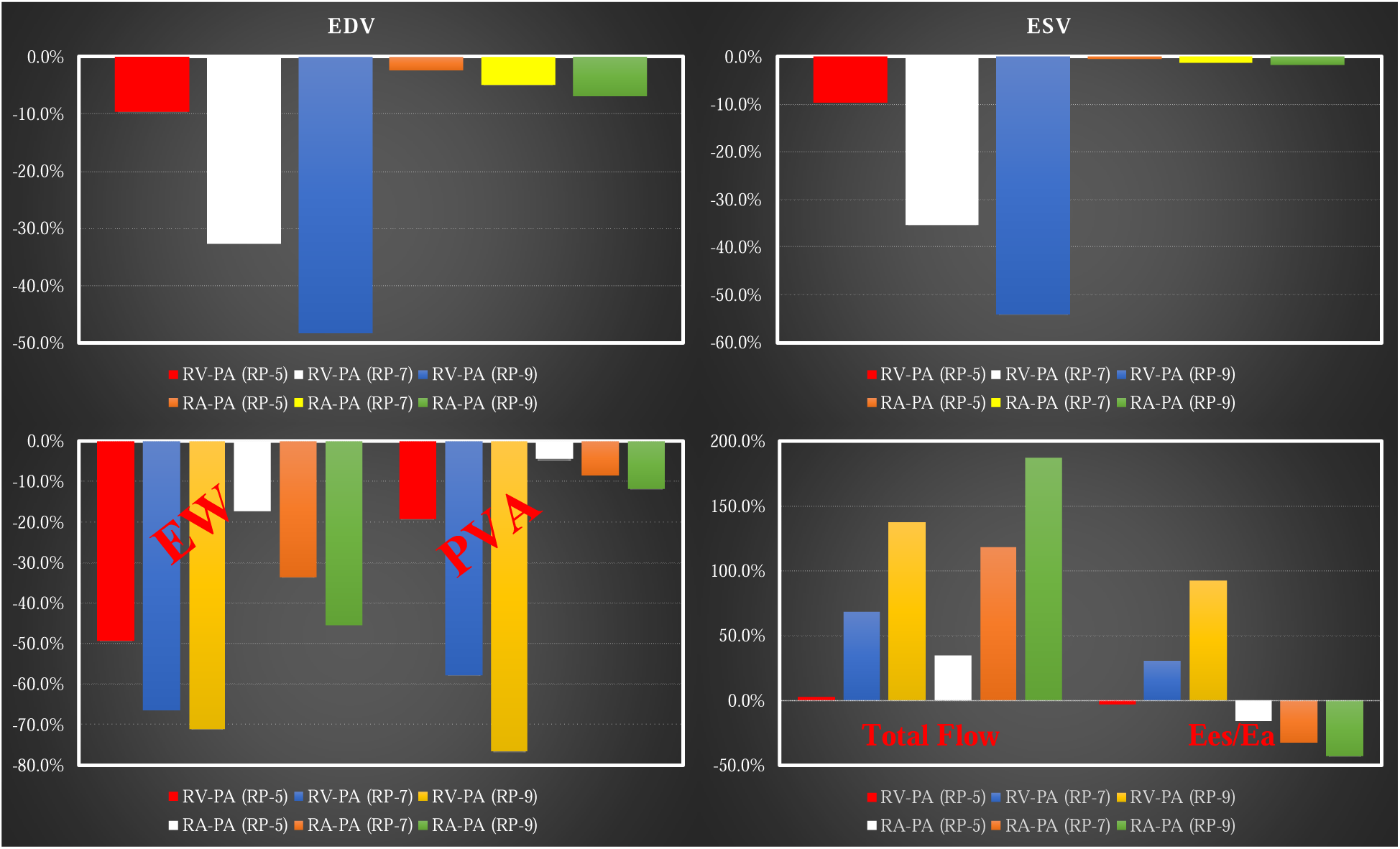
Percentage variations in right EDV (top left panel), right ESV (top right panel), EW and PVA (bottom left panel) and Total Flow with Ees/Ea (bottom right panel) in relation to the following starting conditions: systemic venous pressure (SVP) is kept constant at 10 mmHg; PAWP is set to 10 mmHg. The PAP was kept constant at 60 mmHg. Systemic venous resistance is kept constant at 30 -3 [mmHg‧cm^−3^ sec].

## Discussion

The relationship between structure and function remains an essential element in clinical decision-making, which has to consider how to address the disease and restore normality to a certain degree. The onset of right ventricular dysfunction should be considered in the context of pressure and volume overload or the primary myocardial disease [25, 26]. The management of right heart failure is still difficult because of the complex geometry of the right heart and a lack of specific treatments aimed at recovery of right ventricular function, quite often leading to poor clinical outcome. The use of VV ECMO may have an initial role to play for the treatment of right heart failure although its progression would require a more aggressive treatment such as conversion to VP ECMO using the ProtekDuo, which is a single-site dual-lumen cannula inserted through the right internal jugular vein leading to complete bypass of the right ventricle [15–19] due to RA-PA configuration. Our previous simulation work has confirmed the effective haemodynamic assistance provided by the ProtekDuo as observed in the acute clinical setting [21]. We have also addressed the role of the Impella 2.5 device for left ventricular support either alone or combined with the intra-aortic balloon pump [23, 24]. This has given us the motivation to further investigate the haemodynamic effects of the Impella RP**^®^** device for right ventricular support. The key element of this work is the significantly high correlation between the experimental and simulated pressure-flow curves as shown in Fig. 4, Fig. 5 and Fig. 6. This has given the opportunity to develop realistic simulations for the evaluation of the effects of the Impella RP**^®^**device on a failing right heart. The numerical model implemented within CARDIOSIM^©^ software platform enabled us to study the implications of a possible evolution of the Impella RP^®^ with a view to drain blood from the right ventricle and eject it into the pulmonary artery (RV-PA configuration). The simulation outcome shows that the Impella RP**^®^** device seems to perform better when connected in RV-PA mode with particular reference to unloading of the right ventricle, EW, PVA and Ees/Ea leading to a more adequate ventriculo-arterial coupling. In fact, the RV-PA connection would decrease EW, PVA and Ees/Ea by 38.37%, 21.26% and 18.94% whilst the RA-PA connection would give 22.36%, 9.63% and 20.45% reduction. As far as right ventricular unloading is concerned, the RV-PA connection would reduce EDV and ESV by 18% and 12% whilst the RA-PA connection would give only 9% and 2% reduction. If we consider the right ventricular P-V loop, then a higher leftward shift is observed when the device is connected in RV-PA mode (Fig 9). Percutaneous insertion through the right internal jugular vein would make its use even more appealing given the potential for patient’s mobilisation as observed with the ProtekDuo device. Based on our findings, there is ground for the development of an Impella RP^®^ with the option of RA-PA and RV-PA connection which would make it the device of choice for right ventricular support. Early intervention remains the best option for treating right heart failure and avoiding permanent damage.

## Conclusion

The Impella RP^®^ has the potential to become a valid option for right ventricular support based on current experimental and simulation data. Although already in use, further evaluation in the clinical setting is likely to confirm its potential and lead to a more routinely application for right ventricular support. The potential evolution of the Impella RP^®^ or a similar RVAD, with the option of both RA-PA and RV-PA mode, may lead to the development of a more adaptable device for the assistance of a failing right ventricle.

## Data Availability

All data produced in the present work are contained in the manuscript

https://cardiosim.dsb.cnr.it/

## Acknowledgements

This work was supported by the REMOTE project (grant number F/310117/01/X56) financed by the Italian Ministry of Economic Development.

